# Efficacy and feasibility of Pharmacoscopy-guided treatment for acute myeloid leukemia patients that exhausted all registered therapeutic options

**DOI:** 10.1101/2023.03.28.23287745

**Authors:** Jonas Andreas Schmid, Yasmin Festl, Yannik Severin, Ulrike Bacher, Marie-Noëlle Kronig, Berend Snijder, Thomas Pabst

**Affiliations:** Faculty of Medicine University of Bern, Bern, Switzerland; Department of Biology, Institute of Molecular Systems Biology, ETH Zürich, Zürich, Switzerland; Department of Hematology; Inselspital, University Hospital Bern, University of Bern, Bern, Switzerland; Department of Medical Oncology; Inselspital, University Hospital Bern, University of Bern, Bern, Switzerland

**Keywords:** Acute myeloid leukemia (AML), DARTT-1, *ex vivo* drug screen, personalized treatment, elderly patients, pharmacoscopy, functional precision medicine

## Abstract

Elderly or unfit patients with relapsed acute myeloid leukemia (AML) face a poor prognosis and are likely to rapidly exhaust all registered treatment options. Pharmacoscopy, an image-based *ex vivo* drug screening platform, has previously been suggested as a tool for treatment selection in AML. We used pharmacoscopy to generate personalized treatment recommendations for 30 relapse settings of 24 AML patients which exhausted all standard therapeutic options. We evaluated whether pharmacoscopy can be employed within the narrow timeframe available under an AML relapse setting, how often recommended regimens could be started and whether they provided durable clinical benefits. 17 of 30 screens (56.7%) resulted in patients receiving a recommended therapy, leading to promising trends in clinical response and survival. A drug regimen’s integrated pharmacoscopy score proved to be an excellent predictor of clinical response: Patients receiving a regimen with above-median scores showed significantly higher rates of complete remission (OR 3.01, p < 0.0005) and significantly longer overall survival (ratio 3.39, p < 0.006). We conclude that pharmacoscopy is an efficient and valuable tool for therapy selection in AML at relapse and propose concrete measures to further improve clinical implementation.

## 1. Introduction

Presently, the standard curative treatment strategy for acute myeloid leukemia (AML) is comprised of intensive induction chemotherapy followed by consolidation treatment and allogeneic hematopoietic stem cell transplantation^1^. However, a significant amount of AML patients are unable to undergo such intensive treatment, owing to their advanced age, comorbidities and/or poor performance status^2^. These limitations are even more pronounced in AML patients at relapse due to increased morbidity related to previous treatment attempts, leading to a particularly poor prognosis. Only a few licensed and standardly reimbursed protocols are currently available for AML patients at relapse unable to undergo intensive re-induction protocols and these tend to be quickly exhausted. The situation is aggravated by the rapidly progressive nature of AML at relapse which severely restricts the timeframe to select an optimal therapy regimen^3^. Thus, there is a substantial unmet medical need in this patient group.

The rapid expansion of gene sequencing technology has allowed for the development of molecularly targeted therapy approaches both in newly diagnosed and relapsed AML (e.g., with FLT3 or IDH1/IDH2 mutations). However, such strategies have provided modest clinical benefits and are only open to a relatively small subset of patients at relapse^4,5^. Hence, there currently is a lack of reliable means to select personalized treatment regimens for AML patients at relapse, a challenge that also extends to other cancer entities^6^. Recent studies have demonstrated the effectiveness of an image-based *ex vivo* drug screening platform called pharmacoscopy to retrospectively predict the clinical treatment response in AML patients^7,8^. Furthermore, pharmacoscopy has been shown to provide valuable guidance when selecting targeted therapies in patients with a range of aggressive hematological malignancies lacking standard treatment protocols^9^. Prior studies have tried to integrate *ex vivo* drug screening approaches into the clinical management of AML^7,9,10^ but so far none has focused exclusively on AML patients at relapse that had exhausted all standard therapeutic options, which pose an exceptional challenge due to the severely restricted timeframe available for therapy selection at this stage. Furthermore, previous studies have not addressed the issue of obtaining financial coverage for therapies selected in this way, an issue that is inherent to all currently available drug screening approaches.

In this prospective, non-randomized, single-center observational study, we primarily aim to establish whether pharmacoscopy can be successfully integrated into the clinical decision-making process when selecting individualized treatment protocols for AML patients at relapse that have exhausted all standard therapy options. We evaluated whether the suggested therapies can be made available to the patients within a clinically tolerable time frame and with adequate financial coverage. In addition, we assessed if pharmacoscopy-based therapy recommendations can provide a measurable and lasting clinical benefit in our heavily pretreated and frail study group.

## 2. Materials and Methods

In our prospective, non-randomized, single-center observational study (DARTT-1; NCT05732688; BASEC-ID: 2021-01294, Department of Medical Oncology, University Hospital Bern, Switzerland), we enrolled adult AML patients at relapse who were unable to undergo intensive re-introduction treatments and had exhausted standard treatment options. We excluded patients who had not yet undergone previous treatment attempts or still had other treatment options available. We used an image-based ex vivo drug screening platform, called pharmacoscopy, to generate personalized treatment recommendations for each patient based on samples of peripheral blood, bone marrow, or skin/subcutaneous biopsies. The drug screens were performed at the laboratory of Prof. Berend Snijder, Institute of Molecular Systems Biology, at the ETH Zurich. This group has recently developed and validated a pioneering image-based ex vivo drug screening platform for patients with aggressive hematological malignancies, called pharmacoscopy (Figure 1), and all screens were conducted as previously described ^7–9^. The results of the drug screen were communicated to the treating oncologist in the form of a short list of top-scoring drugs recommended for the treatment of the respective patient, combined with reports on the screening results for all tested compounds. If the recommended compounds could not be made available with adequate financial coverage within a clinically reasonable time frame, the treating oncologist provided the patient with a therapy regimen based on their medical history and previously established in-house guidelines at our department. If the patient chose not to undergo further treatment, they were provided with best supportive care.

**Figure 1.**
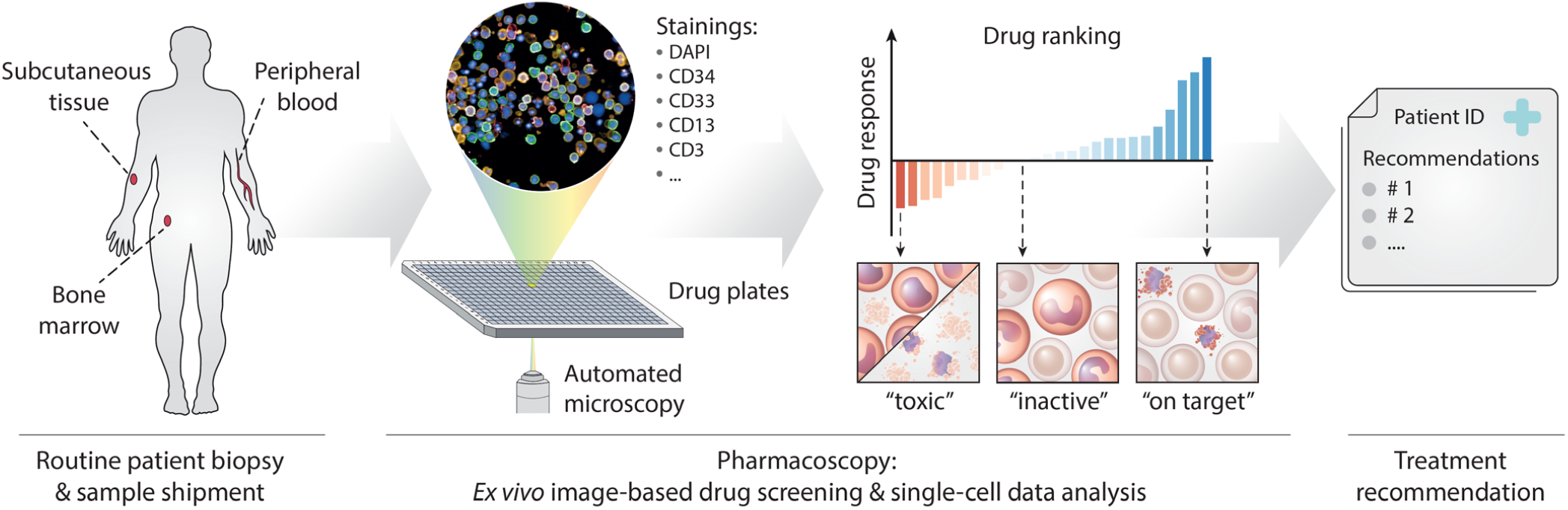
Pharmacoscopy workflow for AML at relapse: Patient samples (bone marrow draws, peripheral blood, or subcutaneous/skin samples) were shipped by courier to the pharmacoscopy lab. There, the cells from the samples were processed by either density centrifugation (blood and bone marrow) or tissue dissociation (skin) and seeded into 384-well imaging plates, with each well containing a chemo- or immunotherapeutic compound from our test library. The plates were then incubated overnight. Immunofluorescence stainings against specific surface antigen characteristics of the patient’s leukemic cells were used to distinguish between healthy cells and malignant blasts. The cells were then subjected to automated confocal microscopy (Opera Phenix, Perkin Elmer) and image analysis using nuclear morphology to quantify the death of malignant and healthy cells, respectively. Based on this readout, the ex-vivo blast reduction capacity (PCY score) was calculated for each compound. Pharmacoscopy reports were provided to the treating oncologists in the form of a short list of top-scoring drugs ranked by their predicted efficacy, as well as complete drug response profiles. The selection of treatment regimens was subsequently based on the pharmacoscopy report and the availability of compounds listed therein. If none of the listed compounds could be made available within a reasonable time frame, therapy regimens were chosen based on previously established in-house guidelines at our department.

We enrolled 24 patients who were screened at least once, and an additional 5 patients were screened a second or third time after relapsing. A total of 30 screening events with subsequent treatment and follow-up were included in our intention to treat population (Figure 2). We obtained signed informed consent from all patients and the study was carried out in accordance with good clinical practice and approved by relevant institutional review boards and regulatory agencies. Screening instance and Patient IDs are not known to anyone besides the authors.

**Figure 2.**
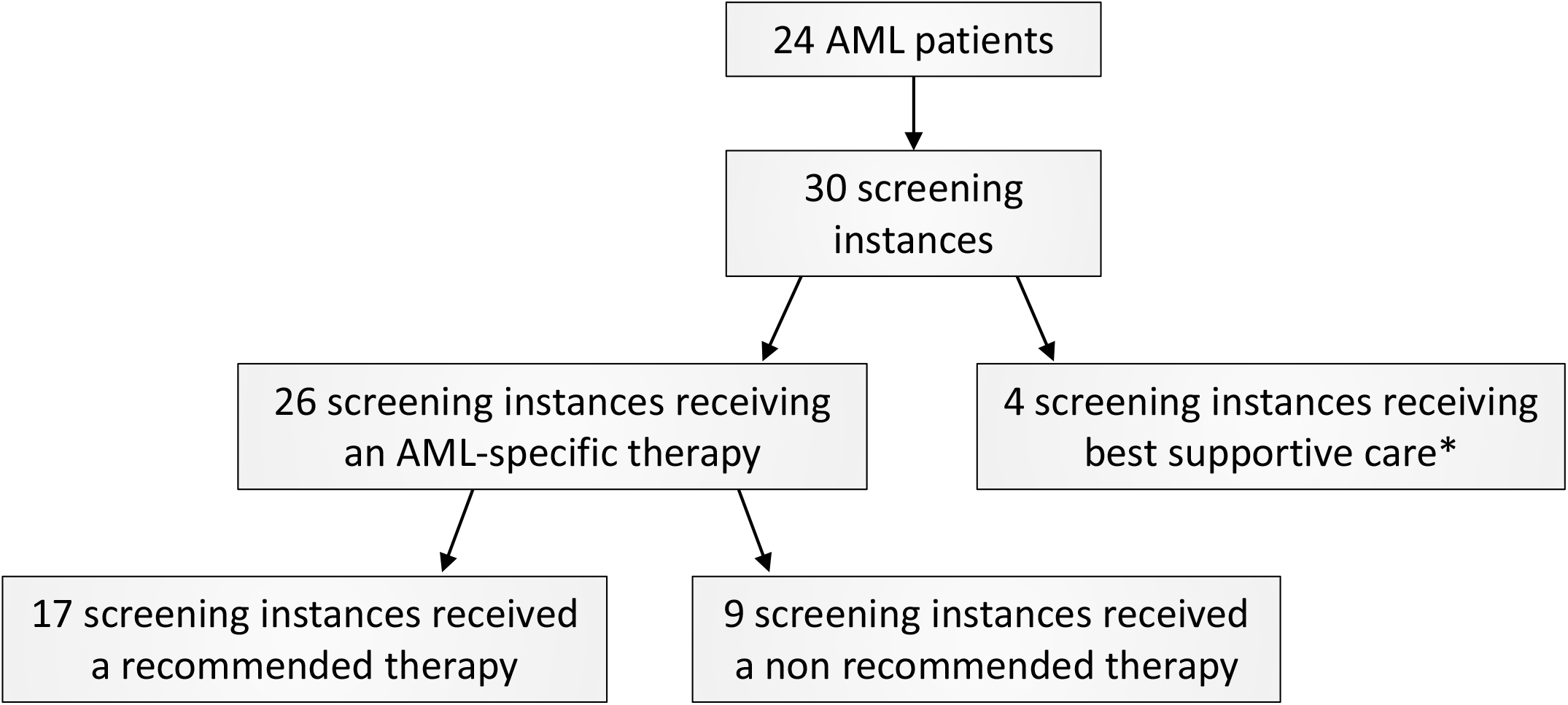
Trial profile: Diagram of study patients. * Patients were switched to best supportive care either because they refused further treatment, or due to their rapidly deteriorating health prohibiting additional AML-specific therapy attempts.

To assess response rates and outcome metrics, we monitored patients using clinical assessments, regular blood testing, and bone marrow aspirations during routine follow-up visits after the start of a newly chosen treatment. We evaluated each screening instance as an individual event and calculated the blast reduction capacity (PCY score, see more detail in supplementary) of a patient’s given therapy regimen by summing up the PCY scores of the individual compounds. Using this metric, we divided patients into two groups based on their therapy regimen’s ex vivo blast reduction capacity (above or below the median of the study population). The primary endpoints of the study were the frequency of patients realizing a complete remission (CR) in the bone marrow, overall survival (OS), and event-free survival (EFS). We assessed these endpoints by comparing patients who received an officially recommended therapy versus those who received a therapy that had not been mentioned in the screening report summary sent to the treating physician. Further details on enrollment, drug screening, statistical analyses and data availability can be found in the extended methods.

## 3 Data availability statement

The data required to reproduce our results are included in the tables and supplementary tables. Additional data generated in this study are not publicly available due to information that could compromise patient privacy or consent but are available upon reasonable request from the corresponding author.

## 4 Results

### 4.1 Patient characteristics

24 patients were enrolled in the study and screened at least once. All patients were pre-treated with a median of 2 previous treatment lines (interquartile range 1-3) and had exhausted all standard therapeutic options for relapsed AML, including all marker-based therapy options. Five out of 24 patients (20.8%) had undergone previous high-dose chemotherapy. The median age of the study population was 68.5 years (interquartile range 66-73) and the gender distribution was 62.5% male vs 37.5% female. According to ELN risk category^1^ 12.5% of the patients were classified as favorable, 37.5% intermediate and 50.0% adverse. The median time from diagnosis to entry into the study was 12.5 months with an interquartile range of 4.5-24.8 months. Demographics and the number of previous treatment lines for these 24 patients are listed in **Table 1**. A more detailed overview of the study population can further be found in **Supplementary Table S2**, listing patient characteristics at the time of first diagnosis.

**Table 1:** Summary of study group. IQR: Interquartilerange. No.: Number

|  |  |
| --- | --- |
| Age at start of the study (years) |  |
| Median (IQR) | 68.5 (66-73) |
| Sex (No. and share of Patients) |  |
| Female | 9.0 (37.5%) |
| Male | 15.0 (62.5%) |
| ELN Risk Category (No. and share of Patients) |  |
| favorable | 3.0 (12.5%) |
| intermediate | 9.0 (37.5%) |
| adverse | 12.0 (50.0%) |
| Time from diagnosis to study (months) |  |
| Median (IQR) | 12.5 (4.5-24.8) |
| Number of previous therapy lines |  |
| Median (IQR) | 2.0 (1.0-3.0) |
| No. and Share of patients with previous high dose chemotherapy | 5.0 (20.8%) |

In the following subsections, we divided our study population according to two stratification strategies. Namely, by whether or not the patient received a therapy regimen recommended by the pharmacoscopy screen and by the *ex vivo* blast reduction capacity of their chosen therapy regimen (i-PCY score of the chosen regimen). Neither of the two patient stratification strategies differed significantly in their patient characteristics (see **Supplementary Table S3**). However, when looking at the number of previous treatment lines, the difference between patients receiving a recommended therapy and those that did not, borders on significance (p 0.0528), with the patients receiving a recommended therapy having a slightly higher median number of previous therapy lines (3 vs 2). In addition to the first pharmacoscopy screen of each patient, five of the 24 patients were screened a second time after a confirmed relapse or progression of the AML under their initially chosen treatment protocol. One of the doubly screened patients was screened a third time after a further relapse, resulting in a total of 30 separate screening events with subsequent therapy choices and clinical follow-up. These 30 screening instances constitute our intention to treat population. Of note, one of the patients presented with a subcutaneous relapse of a bi-specific AML (patient ID 11) for which we screened a skin biopsy, and confirmed the absence of toxicity of the recommended treatments based on an additional screen on a matched bone marrow biopsy that was confirmed AML negative by clinic diagnostics (**Supplementary Figure S1**). See **Supplementary Table S1** for details on each test, including markers used per patient and the pharmacoscopy results for each of the 30 screening instances. All screens were performed during the period between April, 1^st^ 2021 and June, 30^th^, 2022. For the following statistical analyses, each screening event will be treated as a separate data point.

### 4.2 Screening results and treatment choice

**Table 2** lists a summary of the *ex vivo* drug screening results, waiting times, characteristics of treatment choice and duration of therapy administration. The median waiting time for the screening results was 5.0 days (interquartile range 4.0-6.0). A median of 5.5 drugs per patient (interquartile range 4.3-8.8), ranked by their predicted efficacy for the respective patient, were mentioned as official recommendations in the reports sent to the treating physicians. Reports also regularly contained suggestions for potential combinations of the recommended compounds based on the results for the individual components. In 17 (56.7%) instances the drug screen resulted in the patient receiving at least one of the recommended therapy options (either a single drug or a combination, depending on the recommendations of the screen), while no recommended drug was administered in 13 (43.3%) instances. Of these 13 instances, 9 (30.0% of all screening instances) resulted in patients receiving an AML-specific therapy not explicitly recommended by the screen. Four patients did not start another AML-specific therapy due to their rapidly deteriorating condition or refusal of further therapy attempts and were therefore switched to best supportive care (see **Figure 2**). The duration from receiving the drug recommendations to the start of a new therapy regimen was a median of 11 days (IQR 6-24). The median duration of therapy administration was 65.5 days (IQR 26.5-183.8) for all patients receiving an AML specific therapy (recommended and non-recommended). When looking only at patients receiving a regimen recommended by the screen, the median duration of therapy administration was 69.0 days (IQR 21.0-171.0) versus 47.0 days (40.0-214.0) for patients receiving a non-recommended therapy. Out of the 166 single compounds or combinations of compounds in the library used for the *ex vivo* drug screen 59 were recommended (as single compounds and/or in specific combination regimens) at least once in the screening reports (pharmacoscopy reports) sent to the treating oncologist. Out of these, 17 compounds were recommended five times or more. **Table 3** lists the top six most recommended drugs, which were the BCL-2 Inhibitors navitoclax and venetoclax, the alkaloid omacetaxine (formerly named homoharringtonine), the purine analogue cladribine, the proteasome inhibitor carfilzomib and the HDAC-Inhibitor panobinostat. Despite having been among the most frequently recommended drugs, both omacetaxine and panobinostat were never administered to patients in the study due to the difficulty of obtaining financial coverage for their use in AML therapy. Eight individual compounds that had been recommended in a screening report were administered at least once to the respective patient. These include venetoclax, cladribine, cytarabine, azacytidine, navitoclax, carfilzomib, bortezomib and pomalidomide. All the recommended and administered drugs can also be found in the bar plots in **Supplementary Figure S2**.

**Table 2:** Overview of *ex vivo* drug screen and treatment choice. IQR: Interquartilerange, No.: Numbers

|  |  |
| --- | --- |
| Waiting time for results, days |  |
| Median (IQR) | 5.0 (4.0-6.0) |
| No. of recommended compounds per patient |  |
| Median (IQR) | 5.5 (4.3-8.8) |
| Therapy choice per patient (No. of patients and share of intention to treat population) |  |
| Recommendation received | 17.0 (56.7%) |
| Recommendation not received | 13.0 (43.3%) |
| AML specific therapy | 9.0 (30.0%) |
| Best supportive care | 4.0 (13.3%) |
| Duration from recommendation to administration of therapy (days) |  |
| Median (IQR) | 11.0 (6.0-24.0) |
| Duration of therapy administration (days, all patients) |  |
| Median (IQR) | 65.5 (26.5-183.8) |
| Duration of therapy administration (days, recommended therapy) | 69.0 (21.0-171.0) |
| Median (IQR) |  |
| Duration of therapy administration (days, other AML specific therapy) | 47.0 (40.0-214.0) |
| Median (IQR) |  |
| No. of compounds recommended at least once | 59.0 |
| No. of compounds recommended 5 times or more | 17.0 |
| No. of recommended compounds administered to patients at least once | 8.0 |

**Table 3:**
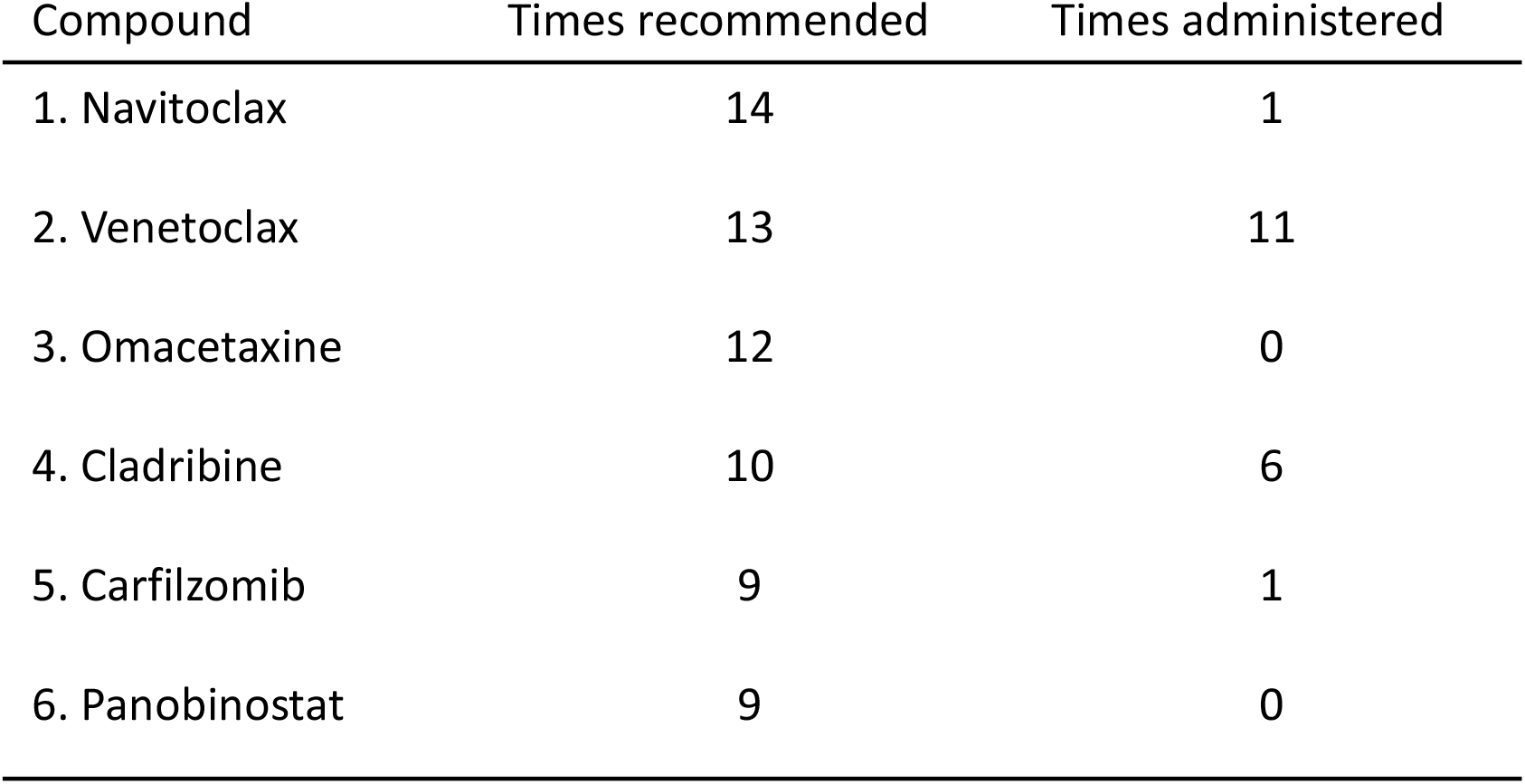
Most frequently recommended drugs. The table lists the 6 most frequently recommended drugs in the screening reports during our study. Despite their frequent recommendation, some of these drugs were never administered to patients due to difficulties in obtaining financial coverage for their use in the treatment of AML patients.

### 4.3 Response

During the study period the overall response to the chosen therapy regimen in the bone marrow could be assessed in 25 screening instances. The results of this analysis are depicted in **Table 4** and **Supplementary Figure S3**. We found that a significantly higher percentage (45.5% vs 21.4%) of patients receiving a drug combination with an above-median *ex vivo* blast reduction capacity (i-PCY score of the chosen regimen) achieved a CR than patients receiving a regimen with a low blast reduction capacity (i-PCY score of the chosen regimen below the median) (OR 3.078, p 0.0005). Conversely, patients that achieved a complete remission tended to receive treatment regimens with a higher *ex vivo* blast reduction capacity (mean i-PCY score 0.278) than patients that did not achieve a CR (mean i-PCY score 0.155) (**Supplementary Figure S4** and **Supplementary Table S4**). When comparing patients receiving a recommended therapy with patients receiving a regimen that had not been officially recommended the difference in the relative number of complete responders is also present albeit less pronounced (35.3% vs 25.0%, OR 1.615, p 0.1646).

**Table 4:**
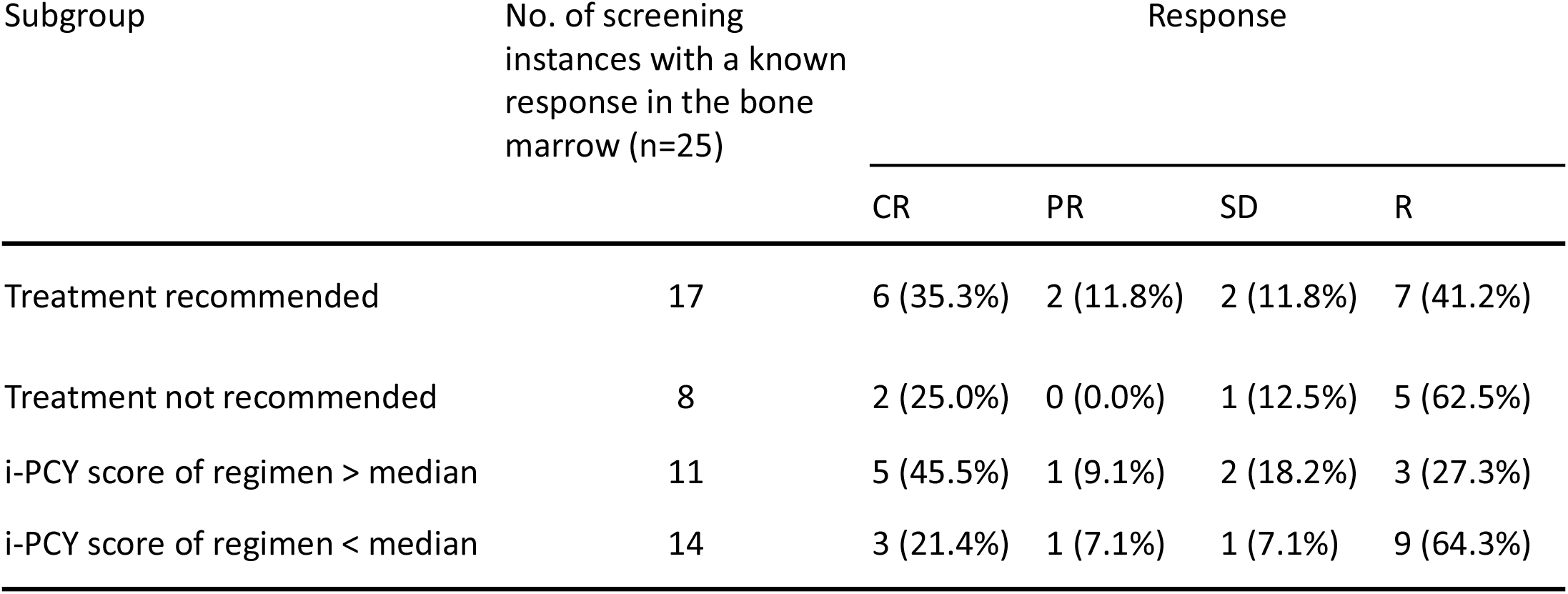
Overall response by subgroups. CR: Complete remission, PR: Partial remission, SD: Stable disease, R: Refractory, No.: Number

| Subgroup | No. of screening instances with a known response in the bone marrow (n=25) | Response |  |  |  |
| --- | --- | --- | --- | --- | --- |
|  |  | CR | PR | SD | R |
| Treatment recommended | 17 | 6 (35.3%) | 2 (11.8%) | 2 (11.8%) | 7 (41.2%) |
| Treatment not recommended | 8 | 2 (25.0%) | 0 (0.0%) | 1 (12.5%) | 5 (62.5%) |
| i-PCY score of regimen > median | 11 | 5 (45.5%) | 1 (9.1%) | 2 (18.2%) | 3 (27.3%) |
| i-PCY score of regimen < median | 14 | 3 (21.4%) | 1 (7.1%) | 1 (7.1%) | 9 (64.3%) |

### 4.4 Outcome

The durability of response to the chosen treatment was evaluated by two different outcome measures consisting of event-free survival (EFS) and overall survival (OS) as defined by standard outcome definitions in AML^1^. Looking at these two outcome measures we found that patients receiving a recommended therapy had a median OS of 18.0 weeks versus 8.0 weeks in the patients not receiving a recommended therapy (ratio 2.250, 95% CI of ratio 1.021 to 4.958) (**Figure 3a**). Median EFS was 11.1 weeks in patients receiving a recommended therapy compared to 6.3 weeks in the group not receiving a recommended regimen (ratio 1.773, 95% CI of ratio 0.805 to 3.906) (**Figure 3b**). When stratifying the intention to treat population by the *ex vivo* blast reduction capacity of their therapy regimen (i-PCY score of the individual regimens) we found that patients receiving a regimen with a blast reduction capacity above the median of the study population showed a median OS of 28.6 weeks as opposed to 8.4 weeks in patients receiving a regimen with low *ex vivo* blast reduction capacity (i-PCY score below the median) (ratio 3.390, 95% CI of ratio 1.506 to 7.632, p 0.006) (**Figure 3c**). Median EFS for patients receiving a regimen with a blast reduction capacity above the median was 12.4 in comparison to 6.4 weeks for the patients treated with a regimen that had a blast reduction capacity below the median (ratio 1.933, 95% CI of ratio 0.859 to 4.353, p 0.0446) (**Figure 3d**). Stratifying only the patients that received an AML-specific therapy by the *ex vivo* blast reduction capacity of their treatment regimen, we found that regimens with an above-median blast reduction capacity led to a median OS of 28.6 weeks versus 11.0 weeks in patients with a treatment regimen scoring below the median (ratio 2.597, 95% CI of ratio 1.062 to 6.354, p 0.0167) (**Figure 3e**). Looking at EFS for the same subgroups we found a median EFS of 12.4 weeks in patients receiving a regimen with an *ex vivo* blast reduction capacity above the median in comparison to 8.1 weeks in patients with a regimen that had a blast reduction capacity below the median (ratio 1.540, 95% CI of ratio 0.629 to 3.767) (**Figure 3f**).

**Figure 3.**
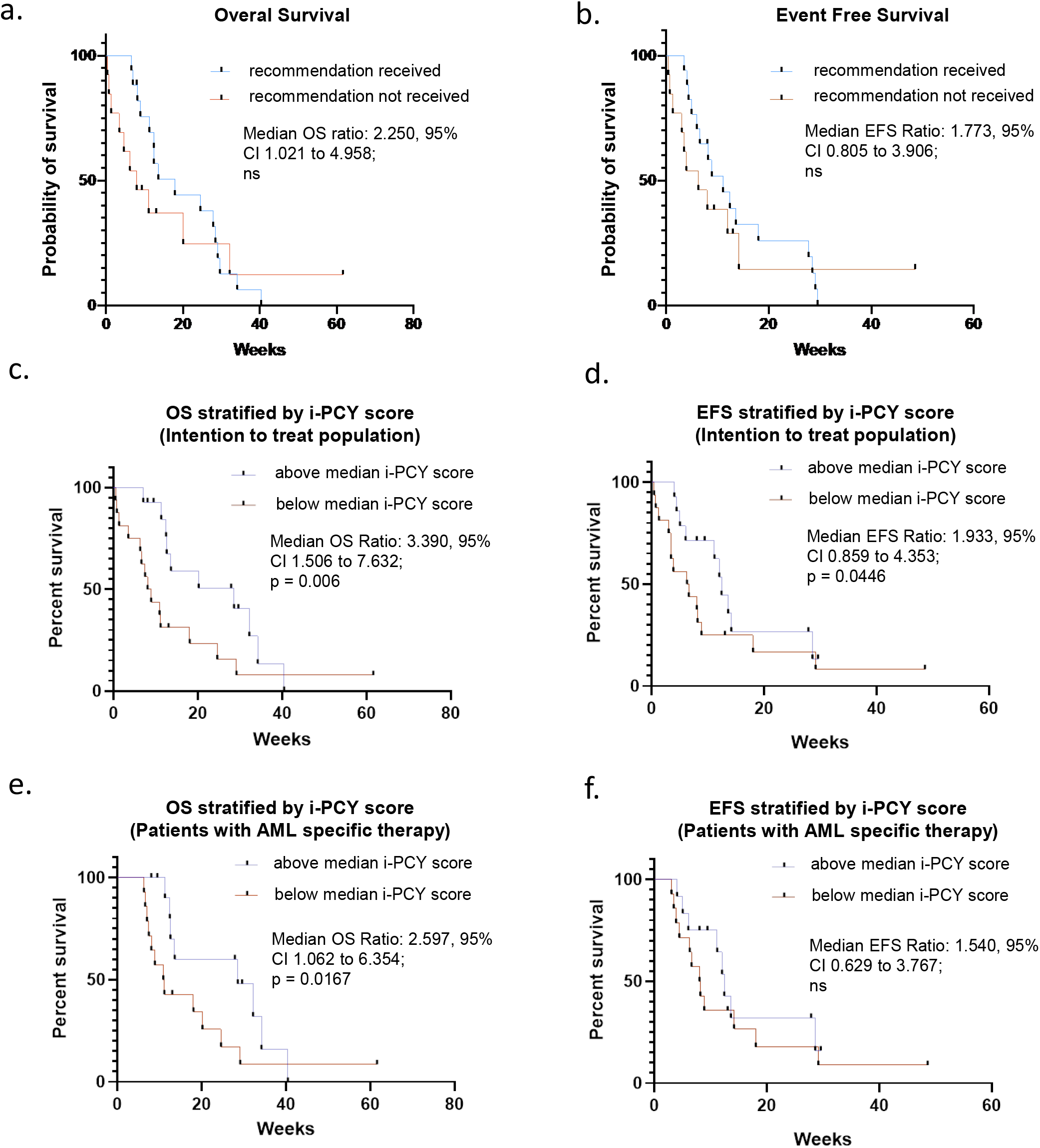
Outcome: **a**. Kaplan-Meier estimates of overall survival in the intention to treat population stratified by whether patients received an officially recommended therapy regimen (17 screening instances) or not (13 screening instances). **b**. Kaplan-Meier estimates of event free survival in the intention to treat population stratified by whether patients received an officially recommended therapy regimen or not (17 vs 13 screening instances). **c**. Kaplan-Meier estimates of overall survival in the intention to treat population stratified by the ex-vivo blast reduction capacity of their respective treatment regimen (i-PCY score) (14 screening instances above median i-PCY score and 16 bellow). **d**. Kaplan-Meier estimates of event free survival in the intention to treat population stratified by the ex-vivo blast reduction capacity of their respective treatment regimen (14 vs 16 screening instances). **e**. Kaplan-Meier estimates of overall survival in patients receiving an AML specific therapy stratified by the ex-vivo blast reduction capacity of their respective treatment regimen (12 screening instances s above median i-PCY score and 14 bellow). **f**. Kaplan-Meier estimates of event free survival in patients receiving an AML specific therapy stratified by the ex-vivo blast reduction capacity of their respective treatment regimen (12 vs 14 screening instances). (OS: Overall survival, EFS: Event free survival). All P values calculated by Gehan-Breslow-Wilcoxon Test.

## 5. Discussion

Despite the expansion of targeted therapies for AML patients, as well as other approaches such as post-transplant maintenance therapy, relapse of AML remains the main cause of mortality in patients with this entity. The exceptionally poor prognosis in this patient group is aggravated further by the very limited availability of registered therapy options at this stage and the rapidly progressive nature of their disease^3^. Thus, improving the prognosis of patients at relapse of AML that have exhausted all registered therapy options remains a critical unsolved issue.

We here evaluated a novel approach, basing treatment decisions in this patient group on a selection of treatments following the recommendations of pharmacoscopy, an automated imaging-based *ex vivo* drug screening platform. Using this platform, we tested chemo-, targeted, and immunotherapeutic drugs for their capacity to selectively eradicate leukemic blasts in patient derived peripheral blood, bone marrow samples or skin/subcutaneous biopsies. In this prospective, non-randomized, single-center observational study, we could demonstrate that the integration of pharmacoscopy into the clinical decision-making for the treatment of AML patients at relapse is technically feasible within the narrow timeframe available and proofs beneficial to the patients.

Patients do not have to undergo strenuous diagnostic tests, as all the material required for the screen can be gained from routine bone marrow punctures, blood draws or skin/subcutaneous biopsies performed during clinical follow-up visits with their treating oncologist. We found that the screening procedure is fast, taking a median of only 5 days to deliver the necessary results and new therapy protocols could be administered within a median of 11 days after receiving the pharmacoscopy results. Thus the procedure does not significantly slow down clinical decision-making, which is a key concern in a rapidly progressing disease such as AML at relapse^1,3^.

In our study cohort, a majority of the intention-to-treat population (56.7%) received a therapy regimen guided by the screening results (either a single drug or a combination, depending on the recommendations of the screen). Patients receiving a regimen recommended by the screen had a median duration of therapy administration of 69.0 days (IQR 21.0-171.0) compared to only 47.0 days (40.0-214.0) for patients receiving a non-recommended therapy. This shows that pharmacoscopy helps direct the choice of therapy regimen and that comprehensive financial coverage could be obtained for the majority of treatment plans derived in this way. Treatments based on official recommendations from the screen appear to be effective, as the rate of CR is higher in patients receiving a recommended therapy, 35.3% versus 25.0% in patients receiving a non-recommended therapy, leading to an odds ratio of 1.615 for a complete remission. This difference in response appears durable with patients receiving a recommended therapy having a 2.3 fold longer OS (95% CI 1.021 to 4.958) and 1.8 fold longer EFS (95% CI 0.805 to 3.906) than those not receiving a recommended regimen. However, we found that the *ex vivo* blast reduction capacity (i-PCY score of a patient’s treatment regimen) is a better ex-post predictor of response and overall outcome than whether the chosen treatment is based on the median 5.5 compounds listed as recommended in the pharmacoscopy reports sent to the treating physician. We found that patients receiving a therapy regimen with an *ex vivo* blast reduction capacity above the median of the study population showed significantly higher relative rates of complete remission in the bone marrow than those receiving a regimen with a blast reduction capacity below that median (OR 3.078, p fisher test 0.0005). Patients receiving regimens with a high *ex vivo* blast reduction capacity (above the population median) also showed significantly longer OS (28.6 weeks vs 8.4 weeks) and EFS (12.4 weeks vs 6.4 weeks). The significant effect of the *ex vivo* blast reduction capacity in a treatment regimen on OS was even present when only looking at patients receiving an AML-specific therapy. Thus, a key takeaway from our study is that when using pharmacoscopy for clinical decision-making, oncologists should base their choice of treatment regimen on the i-PCY score of the respective regimen rather than relying only on the shortlist of recommended compounds. A further finding that strengthens this point is that only a relatively small number of different compounds used in the *ex vivo* drug screen were included in the final treatment regiments of the individual patients. Only eight compounds out of 166 in the screening library were administered to patients at least once after having officially been recommended in the screening reports. These prominently include compounds that have already been established in the treatment of AML such as azacytidine and the BCL-2 inhibitor venetoclax^12^ or cladribine and cytarabine^13^. This skew towards more established treatment regimens is probably owed to their better availability, with health insurance companies being more open to providing financial coverage for more conventional therapy plans. This is an inherent challenge for any drug screening approach using large libraries, since it is not feasible to establish financial coverage for all substances and for every single patient before the actual screens have been run. We, therefore, suggest that future screening reports from pharmacoscopy should always highlight a performance measure of drugs commonly used in the treatment of AML patients. This would allow the treating oncologist to prioritize between readily available standard therapy options even when the highest scoring compounds indicated by the drug screen prove unavailable within a reasonable time frame.

Our study also highlights a number of drugs that have so far not been introduced into standard treatment protocols for AML at relapse but might be promising therapeutic options in the future. The most frequently recommended drug in our screen was the BCL-2 inhibitor navitoclax, a compound that is currently employed in early phase clinical studies for the treatment of solid tumours as well as acute lymphocytic leukemia^14^. The protein translation inhibitor omacetaxine was the third most frequently recommended drug in our screen (behind navitoclax and venetoclax). Omacetaxine has been approved for the treatment of chronic myelogenous leukemia^15^ and has shown promising results in pre-clinical studies on AML treatment^16^. Other frequently recommended drugs include carfilzomib, a drug standardly used in the treatment of multiple myeloma^17^, as well as the HDAC-inhibitor panobinostat which has shown mixed results in both preclinical and early phase clinical studies on AML treatment^18^.

Previous studies have tried to integrate *ex vivo* drug screening into the process of therapy selection in AML patients^9,10^. Malani et al.^10^ reported that 17 out of 29 (58.6%) treated AML patients in their study achieved a PR or better. And, in Kornauth et al.^9^, 6 out of the 14 (42.8%) treated AML patients achieved a PR or better. In comparison with our results, with 8 out of 17 (47.1%) AML patients achieving a PR or better, these prior findings are on par with our results. However, direct comparisons are complicated, as both previous studies included some patients for whom more standard treatment options were available, and considered molecular profiling in parallel to the drug response profiling. In contrast, our patients were all unfit to undergo intensive re-introduction treatments with subsequent hematopoietic stem cell transplantation and had exhausted all available standard treatment protocols, meaning marker-based therapy options were no longer open to our patient cohort. Hence, in contrast to Kornauth et al.^9^ and Malani et al.^10^, we had to rely solely on *ex vivo* drug screening results for therapy selection, making the observed 47.1% PR+CR rate an encouraging signal. Furthermore, in contrast to Malani et al.^10^, we provide a control group consisting of patients for whom a pharmacoscopy guided therapy protocol could not be obtained. And we go beyond these previous studies by addressing the problem of obtaining cost coverage for AML therapy protocols selected on the basis of *ex vivo* drug screens, which is a key issue in the clinical setting.

A major limitation of our study is the lack of randomization of patients into different treatment groups. This flaw is inherent to our study design where it cannot be known whether recommended treatments would prove available for the patient before the screen has been run. However, the significant stratification of the intention-to-treat population by the *ex vivo* blast reduction capacity of their treatment regimen (i-PCY score of the chosen regimen) suggests an efficient design for future randomized trials. Patients could be randomized into an intervention group, which would receive a treatment regimen optimized by the *ex vivo* blast reduction capacity, and into a control group, which would be treated with a regimen according to the physicians choice or in house guidelines.

In conclusion, we can state that pharmacoscopy can rapidly provide valuable decision-making cues for physicians when trying to choose the optimal therapy for AML patients at relapse. The *ex vivo* blast reduction capacity (indicated by a therapy regimens i-PCY score) is a metric that is simple to interpret with the i-PCY score being a numerical value resembling standard laboratory results that physicians interpret on a daily basis. It can readily be used as an indicator to choose between already established treatment protocols or to design novel therapy plans for patients that would otherwise have few suitable treatment options. We suggest that image-based *ex vivo* drug screening may standardly be employed in AML patients at relapse to provide them with optimized treatment plans and further refine the metrics on which physicians base their choice of therapy regimens.

## Contributions

JAS recorded clinical response, carried out statistical analysis, assisted in the enrolment of the patients and wrote the initial draft of the manuscript. YF & YS performed all pharmacoscopy experiments and the associated computational analysis, helped write the final manuscript, and, together with BS, provided treatment recommendations. MNK recruited and enrolled the patients. UB Assisted in conceptualizing the study and writing of the manuscript. BS & TB conceived, supervised, and funded the study, advised on experimental design, data interpretation and helped writing the manuscript. All authors read and approved the final version of the manuscript.

