## Extended materials and methods for "Efficacy and feasibility of Pharmacoscopy-guided treatment for acute myeloid leukemia patients that exhausted all registered therapeutic options"

***Study design and participants***

In our prospective, non-randomized, single-center observational study (DARTT-1; NCT05732688; BASEC-ID: 2021-01294) we enrolled adult patients (≥ aged 18 years) at the Department of Medical Oncology, University Hospital Bern, Switzerland. Included were AML patients at relapse. The inclusion criteria stated that patients needed to be unable to undergo intensive re-introduction treatments with subsequent allogeneic hematopoietic transplantation and must have exhausted all available standard treatment protocols, including all therapy protocols selected on the basis of genetic markers. Conversely, patients were excluded if they had not yet undergone previous treatment attempts, if they still had the possibility of marker-based therapy options or could be submitted to high dose treatment protocols with subsequent allogeneic hematopoietic stem cell transplantation. Peripheral blood, bone marrow samples or skin/subcutaneous biopsies obtained from these patients were subjected to an image-based *ex vivo* drug screening platform, called pharmacoscopy, generating personalized treatment recommendations for each patient. The recommended treatment was administered to the patient if it could be made available with adequate financial coverage within a clinically reasonable time frame. All other patients received the best available treatment based on their medical history and in-house guidelines at our department or were provided with best supportive care if they chose not to undergo further treatment. All patients were then followed closely to determine their response to the chosen treatment and the duration of this response. To be included in the study, the drug screen had to be performed between April 1$^{st}$ 2021 and June 30th 2022. Twenty-four patients were screened at least once. In addition, five patients were screened a second time after relapsing from their initial treatment choice, of which one patient was screened a third time. This yielded a total of 30 screening events at relapse with a subsequent choice of treatment and follow-up, which constitute our intention to treat population (**Figure 1**). The study was carried out in strict accordance with the principles of good clinical practice. Before its start, it was approved by the relevant institutional review boards and regulatory agencies. All patients provided signed informed consent before being enrolled in the study. As of July 1$^{st}$ 2022 the study has been closed and no further enrollment is ongoing at the time of writing.

***Image-based ex vivo drug screening – pharmacoscopy***

The drug screens were performed at the laboratory of Prof. Berend Snijder, Institute of Molecular Systems Biology, at the ETH Zurich. This group has recently developed and validated a pioneering image-based *ex vivo* drug screening platform for patients with aggressive hematological malignancies, called pharmacoscopy, and all screens were conducted as previously described[7–9]. In short, mononuclear cells from bone marrow aspirates, skin/subcutaneous biopsies or peripheral blood were isolated by lymphoprep density centrifugation (Stemcell Technologies) according to manufacturer's instructions and seeded into clear-bottom 384-well drug screening plates, with an average of 5’000 viable patient cells per well. Each well contained a compound or matching control from the previously established drug library. The full drug library contained up to 166 compounds with 113 compounds tested for all patient samples, including both officially approved drugs against hematologic malignancies and other indications as well as licensed study compounds (see **Supplementary Table S1**). All drug plates were prepared by using acoustic, non-contact transfer with the ECHO Liquid Handler (Labcyte). Compounds were tested at 1 and 10µM concentrations, with triplicate repeat wells per concentration, resulting in a total of six wells per drug. After an overnight incubation (at 37° and 5% CO2) with the respective compound, the cells were chemically fixated (1% PFA for 15min at room temperature) and then subjected to immunofluorescence and nuclear staining (DAPI) and imaged by automated confocal microscopy (Opera Phenix, Perkin Elmer) with 20x magnification. The markers used for immunofluorescence were tailored to the specific surface antigen characteristics of the patient’s leukemic cells based on their latest clinical pathology antigen expression assessment reports (see **Supplementary Table S1**), enabling to identify likely malignant from healthy cells in the drug screen. Single-cell image analysis then scored both the identity and viability of each individual cell, as described before[7,11], enabling calculation of a drug score called *pharmacoscopy score* (PCY) for each compound. The PCY score is defined as 1 – (relative fraction of viable target cells in drug-treated conditions / relative fraction of viable target cells under control conditions). If a drug eradicates all target cells without harming non-target cells, the PCY score will be equal to 1. If the drug is killing all non-target cells, the score goes to negative infinity. If a drug destroys both target and non-target cell populations at an equal proportion or has no effect on either population, the score becomes 0. The rationale behind this score is to screen for compounds which specifically kill the malignant target cell population while largely sparing healthy non-target cells[7–9]. The PCY score can also be used to predict the blast reduction capacity of drug combinations by summing up the PCY score values of the individual compounds resulting in the so called integrated pharmacoscopy score (i-PCY)[8]. An overview of the pharmacoscopy workflow for AML patients at relapse is depicted in **Figure 2**.

***Procedures and assessments***

A single sample of peripheral blood, bone marrow or skin/subcutaneous biopsies from each patient, obtained during routinely planned assessments for the diagnostic work-up at relapse, was used for the *ex vivo* screening procedure. For bone marrow samples, 10-12 ml were drawn into heparinized tubes. Blood samples were drawn in a volume of 7.5 ml, using EDTA as an anticoagulant. The samples were shipped the same day by express courier to the pharmacoscopy lab. The results of the screen were communicated to the treating oncologist in the form of a short list of top-scoring drugs recommended for the treatment of the respective patient, combined with PDF reports on the screening results for all tested compounds. The oncologist then tried to make the recommended compounds (either single compounds or drug combinations) available to the patient by applying to insurance companies or drug manufacturers for cost coverage. If multiple recommended compounds were combined in one treatment regimen, the combinations were chosen based on known toxicities and the patents previous clinical history. If the cost coverage for the recommended compounds could not be achieved within a clinically reasonable time, the treating oncologist provided the patient with a therapy regimen based on their medical history and previously established in-house guidelines at our department. If the patient chose not to undergo further treatment, they were provided with best supportive care. The patients were monitored using clinical assessments, regular blood testing and bone marrow aspirations performed during routine follow-up visits after the start of a newly chosen treatment. If a patient relapsed again or showed progressive disease under a chosen treatment regimen within the timeframe of the study, they were subjected to an additional round of *ex vivo* drug screening followed by the selection of another regimen and subsequent clinical follow-up. Each screening instance was treated as an individual event and evaluated separately for the statistical analysis of response rates and outcome metrics. The primary endpoints of the study were the frequency of patients realizing a complete remission (CR) in the bone marrow, overall survival (OS) and event-free survival (EFS), assessed following standard outcome measures for clinical trials in AML[1]. To appraise these endpoints, the patients were divided into four none mutually exclusive subgroups. The first two subgroups compared against each other consisted of patients that received an officially recommended therapy versus patients receiving a therapy that had not officially been mentioned in the screening report summary sent to the treating physician. The other two subgroups were derived ex-post by calculating the i-PCY score (by summing up the PCY scores of the individual compounds), which is an indicator for the blast reduction capacity of a patient's given therapy regimen. Using this metric, we could divide patients into an analysis group that received treatment regimens with a higher than average *ex vivo* blast reduction capacity (i-PCY score above the median of the study population) and a group receiving therapy protocols with a lower blast reduction capacity (i-PCY score below the median).

***Statistical analyses***

The clinical data cutoff date for our study was August 1st 2022 and no further patients were enrolled into the study. The intention to treat population consisted of 24 patients comprising 30 individual screening events. The distribution of overall and event-free survival in the different subgroups was estimated using the Kaplan-Meier method and p-values for the survival analysis were calculated with the Gehan-Breslow-Wilcoxon Test. To calculate the p-values for the difference in the relative rate of CR between the different subgroups we used Fisher's exact test, and the respective odds ratios (OR) were calculated according to the Baptista-Pike method. To calculate the p-values for the demographic differences between our patient stratification strategies we used Pearson's Chi-squared test with Yates' continuity correction and unpaired two-samples T-test. The data were analyzed using Graphpad Prism® Version 8.0.1 (Graphpad Software Inc., La Jolla, CA) and R version 3.1.2 (The R Foundation for Statistical Computing).
