## Supplementary Figure 1 for "Efficacy and feasibility of Pharmacoscopy-guided treatment for acute myeloid leukemia patients that exhausted all registered therapeutic options"

a.

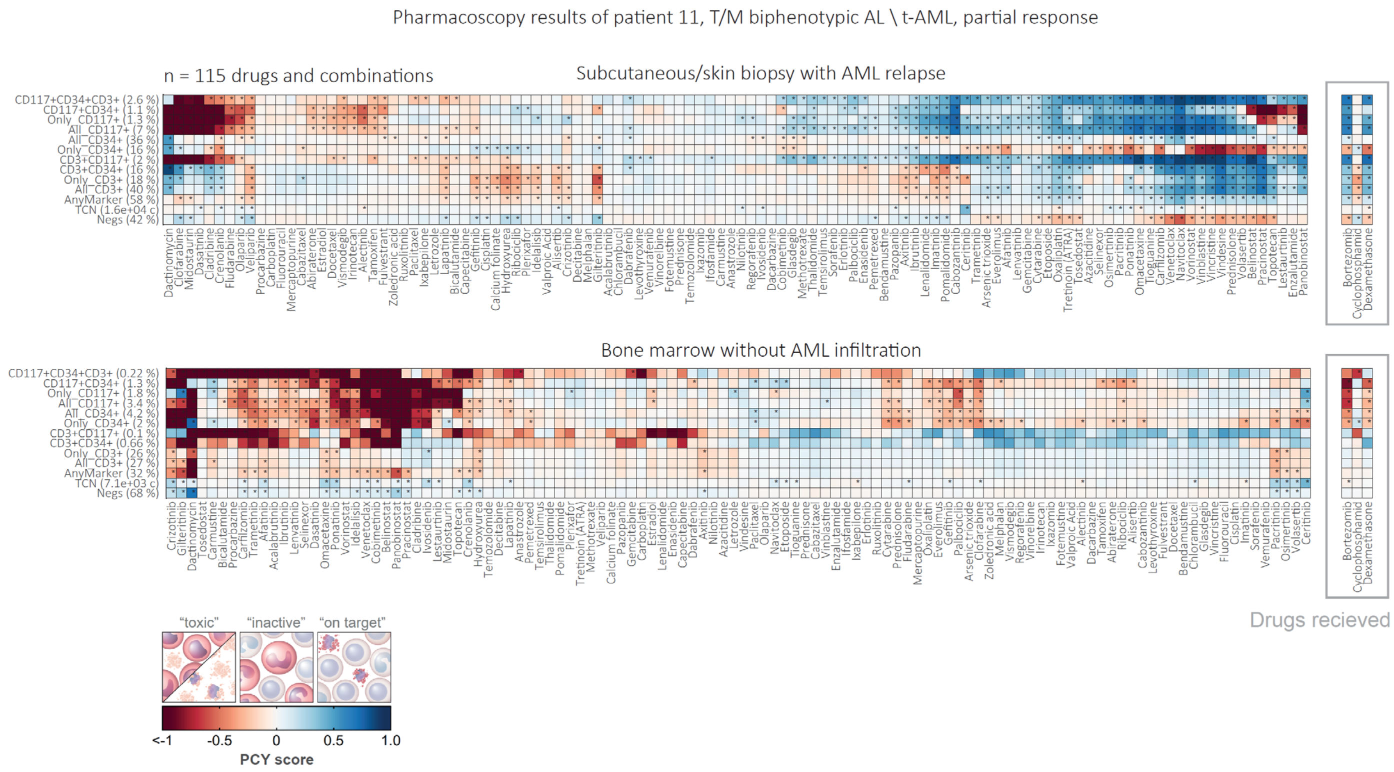


**Supplementary Figure S1. Patient-matched drug testing in skin biopsy (with AML infiltration) and bone marrow** (without AML infiltration)**:** Patient 11 presented at relapse with a subcutaneous AML, but without AML cells detected either in peripheral blood or bone marrow. For this patient, we performed the drug test and treatment identification based on the skin biopsy (upper panel), and confirmed the absence of toxicity for recommended drugs in a matching – but confirmed negative – bone marrow biopsy (lower panel). Inserts on the right show the results for the received therapy. Colors indicate the relative target fraction. Stars indicate significance at $p < 0.05$ by Student's T test.
