## Supplementary Figure 2 for "Efficacy and feasibility of Pharmacoscopy-guided treatment for acute myeloid leukemia patients that exhausted all registered therapeutic options"

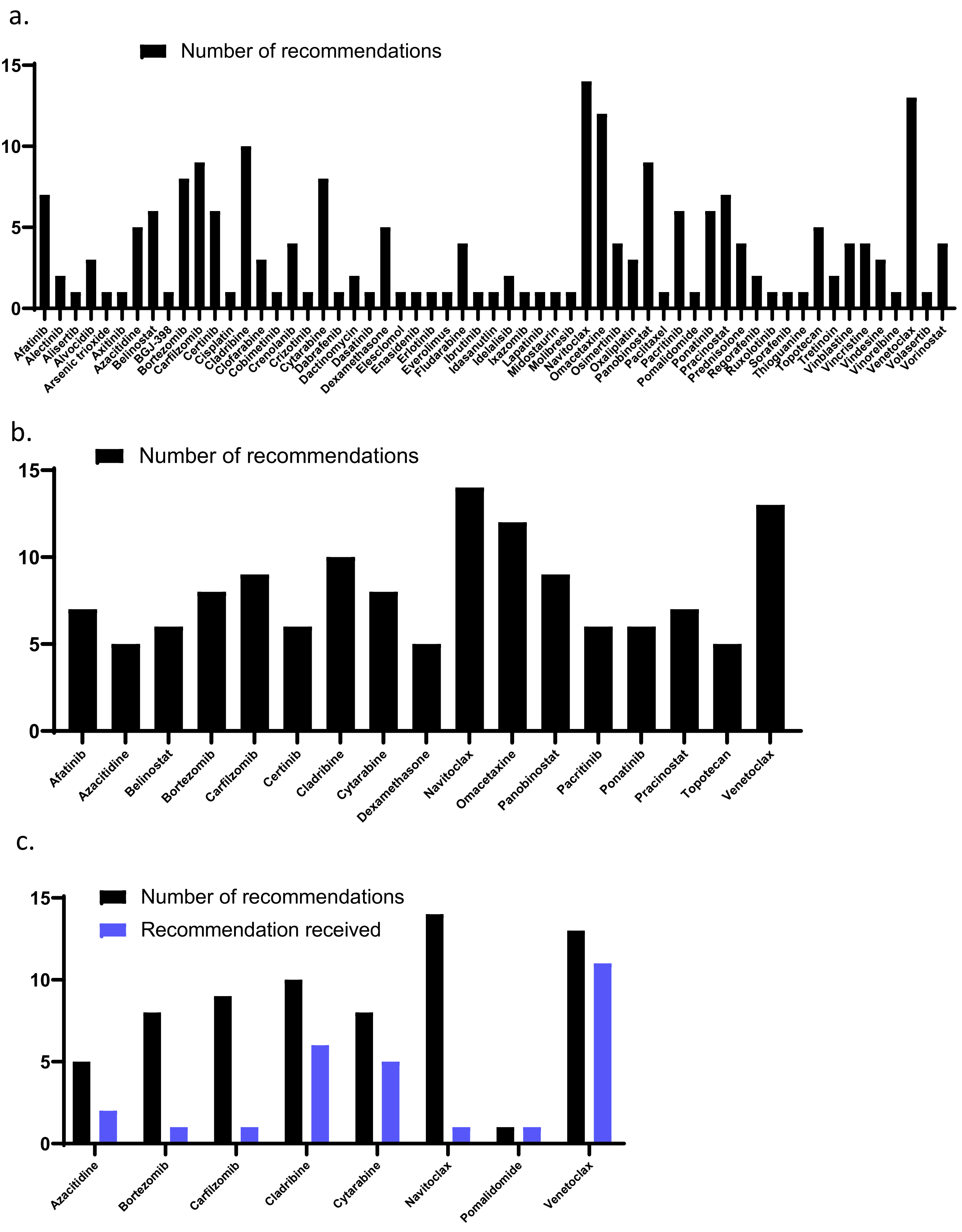


**Supplementary Figure S2. Recommended drugs: a.** drugs that were officially recommended in at least one of the screening instances (59 compounds out of 166 in the screening library). **b.** drugs that were recommended in 5 or more screening instances (17 compounds out of 166 in the screening library). **c.** recommended drugs that were included in a treatment regimen administered to the respective patients (8 compounds out of 166 in the screening library).
