## Supplementary Figure 3 for "Efficacy and feasibility of Pharmacoscopy-guided treatment for acute myeloid leukemia patients that exhausted all registered therapeutic options"

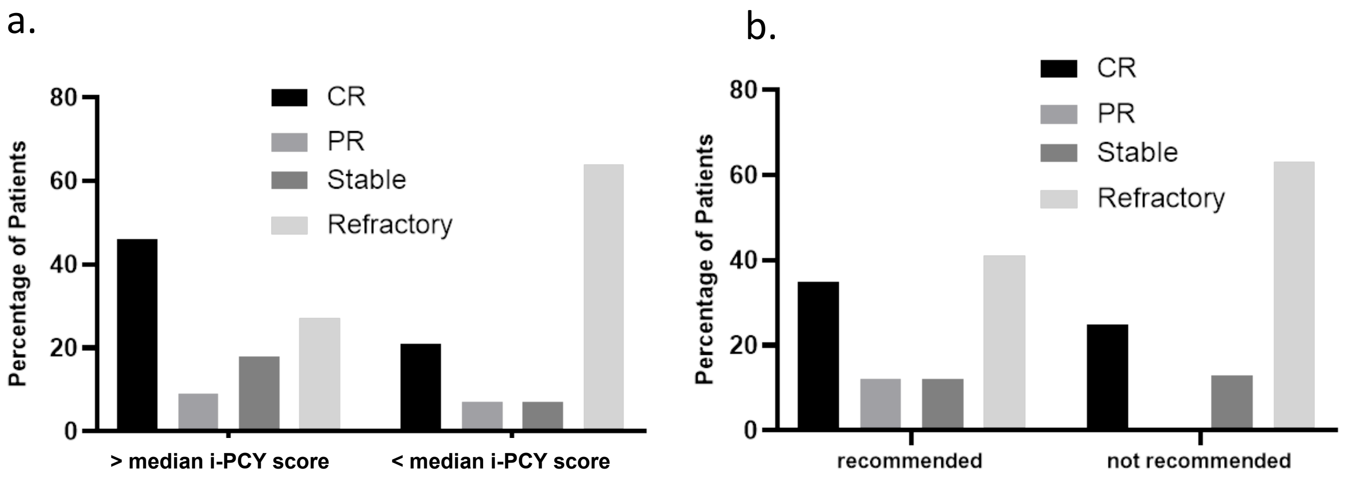


**Supplementary Figure S3: Response stratified by i-PCY score: a.** percentage of screening instances receiving an AML specific therapy and achieving different degrees of response in the bone marrow stratified by the ex-vivo blast reduction capacity (i-PCY score of the chosen regimen) of their respective treatment regimen (11 screening instances with > median i-PCY score and 14 < median i-PCY score). **b.** percentage of screening instances receiving an AML specific therapy and achieving different degrees of response in the bone marrow stratified by whether they received an officially recommended therapy regimen or not (17 screening instances in the recommended group and 8 in the not recommended). (CR: Complete remission, PR: Partial remission, Stable: Stable disease, Refractory: Refractory disease).

Response in the bone marrow could be determined for 25 screening instances. The data depicted in this figure is also included in table 4 of the main manuscript.
