## Supplementary Figure 4 for "Efficacy and feasibility of Pharmacoscopy-guided treatment for acute myeloid leukemia patients that exhausted all registered therapeutic options"

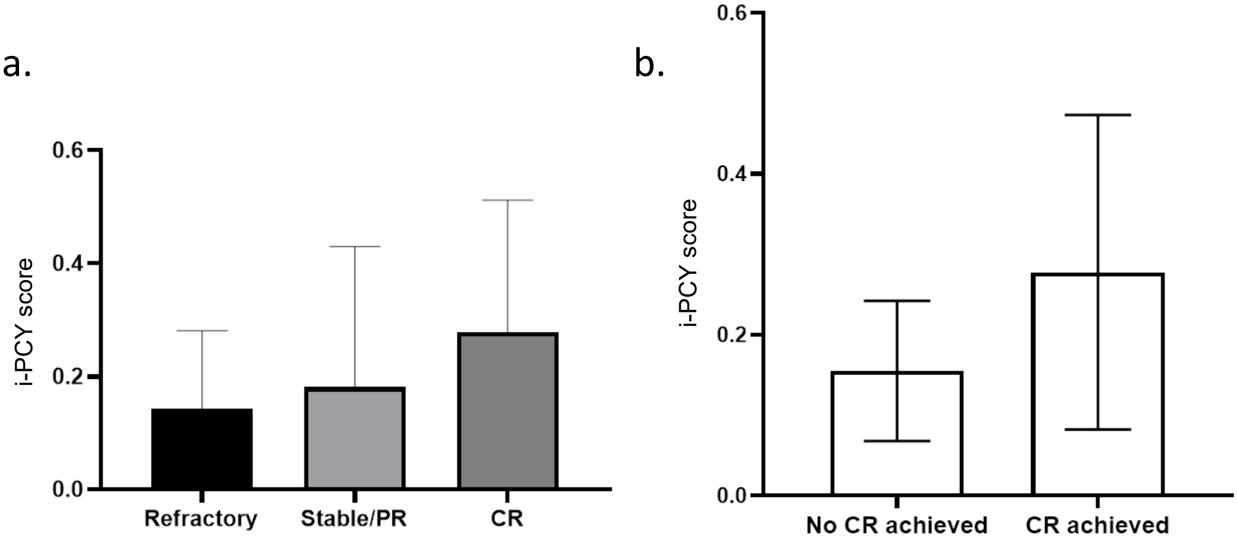


**Supplementary Figure S4. i-PCY scroe of treatment regimen stratified by response: a.** distribution of i-PCY score of therapy regimens (ex-vivo blast reduction capacity) for screening instances that received an AML specific therapy achieving different degrees of response in the bone marrow (8 screening instances achieved a CR, 5 Stable/PR and 12 were refractory). The bar plots show the average i-PCY score for each response category as well as the standard deviation. **b.** percentage of screening instances receiving an AML specific therapy and achieving either a complete remission (8 screening instances) or no complete remission (17 screening instances). The bar plot shows the average i-PCY score(ex-vivo blast reduction capacity) for each response category as well as the standard deviation (CR: Complete remission, PR: Partial remission, Stable: Stable disease, Refractory: Refractory disease)
Response in the bone marrow could be determined for 25 screening instances. The data depicted in this figure is also included in table 4 of the main manuscript and in supplementary table S4.
